# Objective sensory testing methods reveal a higher prevalence of olfactory loss in COVID-19–positive patients compared to subjective methods: A systematic review and meta-analysis

**DOI:** 10.1101/2020.07.04.20145870

**Authors:** Mackenzie E. Hannum, Vicente A. Ramirez, Sarah J. Lipson, Riley D. Herriman, Aurora K. Toskala, Cailu Lin, Paule V. Joseph, Danielle R. Reed

**Author notes:** Please address correspondence: Danielle R. Reed, Ph.D., Monell Chemical Senses Center, Philadelphia PA 19104. These authors contributed equally to this work (Mackenzie E. Hannum, Vicente A. Ramirez).

## Abstract

Severe acute respiratory syndrome coronavirus 2 (SARS-CoV-2), which causes coronavirus disease 2019 (COVID-19), has currently infected over 6.5 million people worldwide. In response to the pandemic, numerous studies have tried to identify causes and symptoms of the disease. Emerging evidence supports recently acquired anosmia (complete loss of smell) and hyposmia (partial loss of smell) as symptoms of COVID-19, but studies of olfactory dysfunction show a wide range of prevalence, from 5% to 98%. We undertook a search of Pubmed/Medline and Google Scholar with the keywords “COVID-19,” “smell,” and/or “olfaction.” We included any study that quantified olfactory loss as a symptom of COVID-19. Studies were grouped and compared based on the type of method used to measure smell loss—subjective measures such as self-reported smell loss versus objective measures using rated stimuli—to determine if prevalence rate differed by method type. For each study, 95% confidence intervals (CIs) were calculated from point estimates of olfactory disturbance rates. We identified 34 articles quantifying anosmia as a symptom of COVID-19, collected from cases identified from January 16 to April 30, 2020. The pooled prevalence estimate of smell loss was 77% when assessed through objective measurements (95% CI of 61.4-89.2%) and 45% with subjective measurements (95% CI of 31.1-58.5%). Objective measures are a more sensitive method to identify smell loss as a result of infection with SARS-CoV-2; the use of subjective measures, while expedient during the early stages of the pandemic, underestimates the true prevalence of smell loss.

## Introduction

In December 2019, an outbreak of a novel coronavirus disease, coronavirus disease 2019 (COVID-19), caused by severe acute respiratory syndrome coronavirus 2 (SARS-CoV-2) that originated in Wuhan, China, rapidly spread to almost every country worldwide. As of June 5, 2020, over 6.5 million cases have been identified and over 387,155 deaths have been attributed to the virus (1). The most common symptoms of infection include fever, dry cough, and fatigue (1). Other accepted symptoms include difficulty breathing, sore throat, headache, nasal congestion, diarrhea, skin rash, and body aches and pains (1, 2). However, as knowledge about the virus increased with more confirmed cases, reports of loss of smell and/or taste started to arise. Other the past few months, COVID-19 research has investigated olfactory and taste disturbances as potential symptoms of COVID-19 (3-5). Many of these disturbances include the immediate onset of a complete loss of smell (anosmia) and/or taste (ageusia); other studies report hyposmia, a reduction in perceived odor intensity. Therefore, the Centers for Disease Control and Prevention and the World Health Organization officially included losses of smell and taste as symptoms of COVID-19, though less prevalent than some other symptoms (1).

While olfactory loss is a common symptom of numerous viral respiratory infections (6), recent reports suggest its prevalence rate might be higher with SARs-CoV-2 infection (7). However, there is a wide reported range of olfactory disturbance prevalence, from 5% (8) to 98% (9). Thus, there is a need to better quantify smell loss during the COVID-19 pandemic (10). Differences in the reported values may be attributed to different recruiting and sampling methodologies, the range of symptom severity across patients, and the amount of information about COVID-19 available at the time of data collection (e.g., symptom recognition).

However, different data collection techniques used by researchers and health care professionals might also account for the different prevalence rates reported. There are two general types of methods to measure smell loss: objective and subjective. Objective measures of smell encompass psychophysical testing designed to measure and quantify human responses to physical stimuli. Though sparsely used in COVID-19 research to date, current psychophysical techniques encompass odor threshold tests to determine the lowest concentration of an odor that can be detected, odor discrimination tests to measure the ability to differentiate between odors, and odor identification tests, assessing the ability to correctly name odor qualities. When possible, these tests are performed repeatedly over several days to measure changes in a patient’s smell abilities over time. Historically, these objective tests are often executed in a laboratory setting, under surveillance of a researcher or health care professional, to ensure proper completion. Examples of odor threshold tests in a COVID-19 population involve the use of butanol or phenylethyl alcohol at different concentrations (11-13). The Sniffin’ Sticks test, an odor discrimination and threshold test, is another method to quantify human olfactory performance (14) used now in COVID-19 patients (15, 16). However, due to the global presence of stay-at-home orders, many researchers have adapted these objective methods to enable testing at home, by patients themselves, with common household odorants (5, 11).

A more common technique employed to quantify smell loss in the COVID-19 population uses subjective methods, self-report through patient questionnaires or interview or the extraction of symptomatic information from a patient’s electronic health records (8, 17, 18). However, collecting information from records can be prone to underestimation of smell loss due to an initial lack of awareness that it is a symptom of COVID-19. Other subjective methods directly ask patients about their own perceived sense of smell through an online questionnaire (7, 19), over the phone (18), or in person with a doctor (20, 21). However, retrospective assessments through self-report measures are often prone to recall bias (22). The present review provides a comprehensive assessment of methodologies currently employed to quantify smell loss in COVID-19–positive patients and examines whether method type affects reported prevalence of smell loss in COVID-19 patients. Another recent systematic review examined the prevalence of olfactory loss as a symptom in COVID-19; however, it contained data collected up until April 19, 2020, encompassed different inclusion criteria, included only 10 papers in its analysis, and additionally examined gustatory dysfunction (23). Building on that prior meta-analysis, we sought to compare differences in prevalence rates of smell loss collected via objective versus subjective methods. We included any study that quantified smell loss as a symptom of COVID-19, summarizing reports available up until June 5, 2020.

## Methods

### Article Selection

This systematic review and meta-analysis followed the PRISMA (Preferred Reporting Items for Systematic Reviews and Meta-Analyses) guidelines (24). Figure 1 outlines the steps taken to select articles for inclusion in the meta-analysis. First, Pubmed/Medline and Google Scholar were used to retrieve literature with the keyword “COVID-19” plus “smell” and/or “olfaction” on May 15, 2020, and manual search of relevant articles via Google Scholar was also performed on June 4, 2020, yielding a total of 78 articles.

**Figure 1.**
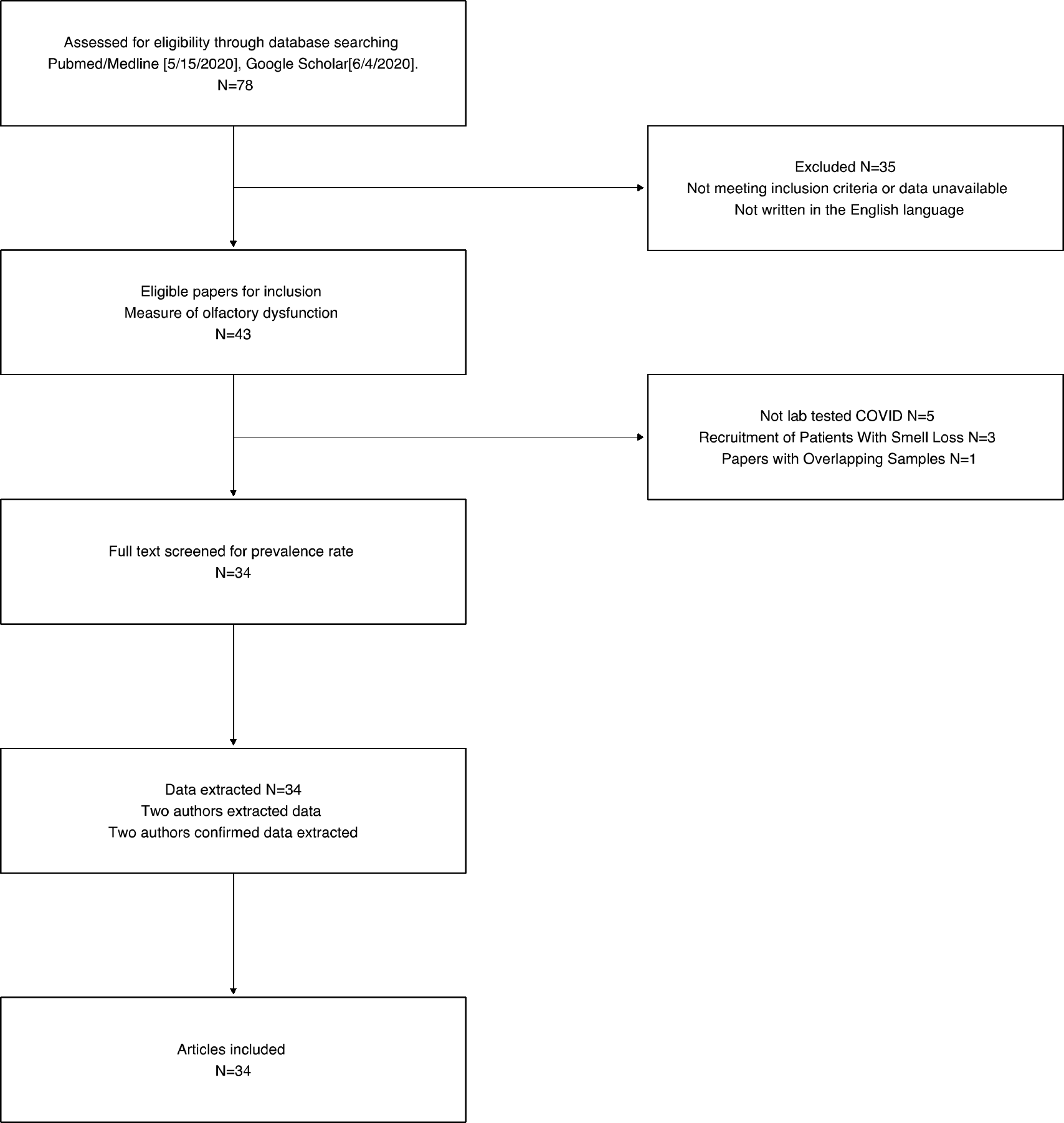
CONSORT flow diagram of the selection process for articles included in this meta-analysis.

Titles and abstracts were then screened for their relevance to the topic. Thirty-two articles were initially excluded during the screening test if they were not about smell loss and COVID19, did not report cases or percentage of patients with smell loss, or if they were not written In the English language. If an abstract referenced a measure of prevalence of olfactory dysfunction in COVID-19–positive patients, it was included in the pool of articles (*n* = 43). Full texts were then screened to confirm positive identification of COVID-19 in patients via a nasopharyngeal swab, throat swab, RT-PCR–confirmed laboratory test, or a clinical assessment by medical professional. Five articles were excluded because patients had not tested positive for COVID-19 by one of these methods (25-29). Three articles were excluded due to population bias, with patients recruited for a specific symptom alone (e.g., olfactory disorders) (30-32). The exact data needed for our analysis were not reported in three articles, which were thus excluded (15, 33, 34). Lastly, one article was excluded (4) because of potential data overlap with another report by the same author (35). A total of 34 papers were included in the meta-analysis.

Prevalence rate of olfactory loss in COVID-19 patients was then extracted as the number of reported cases with olfactory loss divided by the total population of COVID-19 patients surveyed. An exception was made for articles that reported taste and/or smell dysfunction when anosmia or hyposmia were not specifically reported. Articles were also labeled as using either objective or subjective methods to measure smell loss. Studies having patients smell a substance, including both household items being self-administered in their own home and smelling items in a laboratory setting, were classified as objective measures. All other methods, for example, self-reports of overall smell loss, were considered subjective measures.

Due to differences in how data were specifically collected across the studies, further inclusion restrictions regarding how smell loss was reported were established. When smell loss was reported in tandem with taste loss (e.g., “loss of taste or smell”), this value was extracted. If smell and taste loss were reported together as well as separately, both values of positive cases with smell loss symptoms were summed to represent all patients presenting smell loss; these values did not include overlapping patients. If smell and taste loss were reported separately, smell-loss-only values were included.

Three authors (RDH, AKT, and VAR) performed the initial screen and data extraction, and two additional authors (MEH and SJL) validated and resolved disagreements in the data extracted from the articles.

### Risk-of-Bias Assessment

Quality of the articles selected was analyzed with a risk-of-bias assessment checklist adapted from Hoy et al (36). Risk-of-bias assessment was completed by two authors (RDH and SJL) using an assessment tool outlined by Hoy et al. (36), as described and adapted by Tong et al (23). Any differences were resolved by two additional authors (VAR and AKT). The nine criteria are detailed in Supplementary Table S1. Specific questions were scored as 0 (No) or 1 (Yes) for each item, with summary scores of low (0-3), moderate (4-6), and high (7-9) risk of bias for the entire study. Supplementary Table S1 contains the full risk-of-bias assessment for each article.

### Statistical Analysis

All statistical analyses were performed using R 3.6.0 (37) and RStudio 1.2.1564 (38). Point estimates of the prevalence rate of olfactory dysfunction were made by dividing the proportion of cases of olfactory loss by the total number of subjects included in the study. Studies were categorized into two subgroups based on the methodologies employed (objective versus subjective) for additional analysis. A 95% CI was calculated using the Wilson score estimate of the confidence interval, a robust method that is reliable across small and large sample sizes (39).

Three sets of pooled prevalence rates were then computed and reported for both a fixed-effect model and a random-effect model. In the fixed-effect model, we assume that there is one true effect size that underlies all the studies in this analysis and that all differences in observed effects are due to sampling error. In the random-effects model, we allow that the true effect size might differ among studies. An overall pooled prevalence rate was computed for all 34 studies to determine overall prevalence of smell loss in COVID-19 patients, and pooled prevalence rates for the two subgroups: objective methodologies (*N*=6 studies) and subjective methodologies (*N*=28 studies). Pooled prevalence rates were calculated using the *meta* package in R (40). An inverse variance approach was used to approximate the weighting scheme between studies, and the Freeman-Tukey double arcsine transformation was used in calculating the weighting scheme. Heterogeneity was assessed using Cochran’s *Q* and *I*^2^. Tests for heterogeneity were cut off at Cochran *Q*-values that were significant (*p* < 0.05) and *I*^2^ > 50%, because an *I*^2^ of 30-50% was suggested as a cutoff for moderate heterogeneity by Higgins and Thompson (41). A random-effects model was used to account for instances of high heterogeneity between studies and to provide a conservative prevalence estimate. The R scripts and compiled data used for this analysis are available without restriction on GitHub (https://github.com/vramirez4/COVID19-OlfactoryLoss).

## Results

### Study characteristics

Thirty-four studies were included in this meta-analysis, encompassing data collected from January 16, 2020, to April 30, 2020. Table 1 summarizes relevant details from the articles. Figure 2 lists *n*/*N*-values (events/total) for each study. All studies examined COVID-19–positive patients, though the levels of symptom severity, settings (hospitalized or home quarantine), and dates of infection differed across the studies and were not controlled in this meta-analysis. Furthermore, data were collected from around the world, which could increase heterogeneity across the studies. Six studies were classified as using objective methodologies: they measured smell loss in COVID-19 patients by calculating their odor threshold sensitivity, odor discrimination ability and/or odor identification ability with actual odorants, either at home or in hospital settings. Twenty-eight studies were classified as using subjective methodologies: they measured smell loss via questionnaires, surveys, and interviews.

**Table 1.**
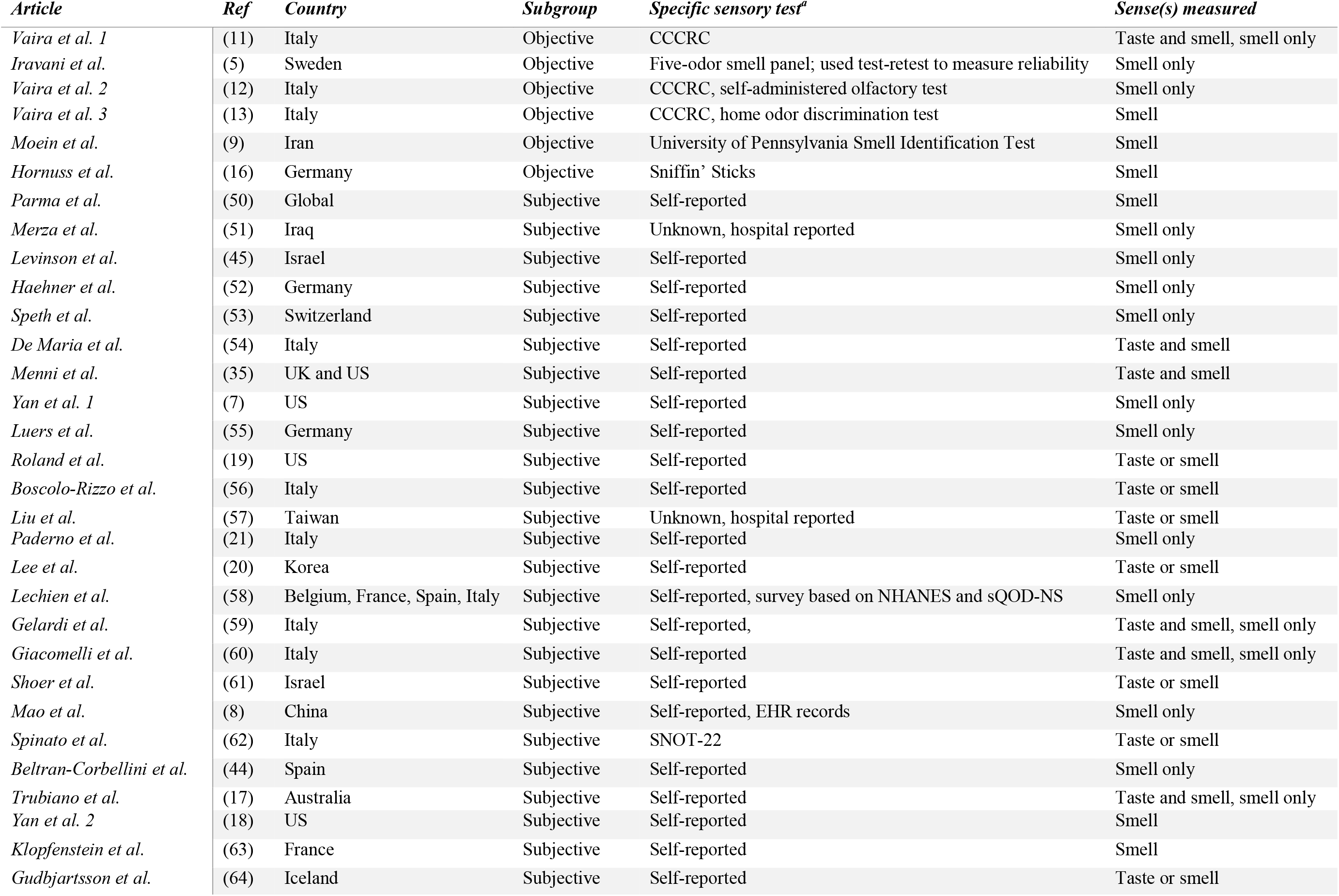
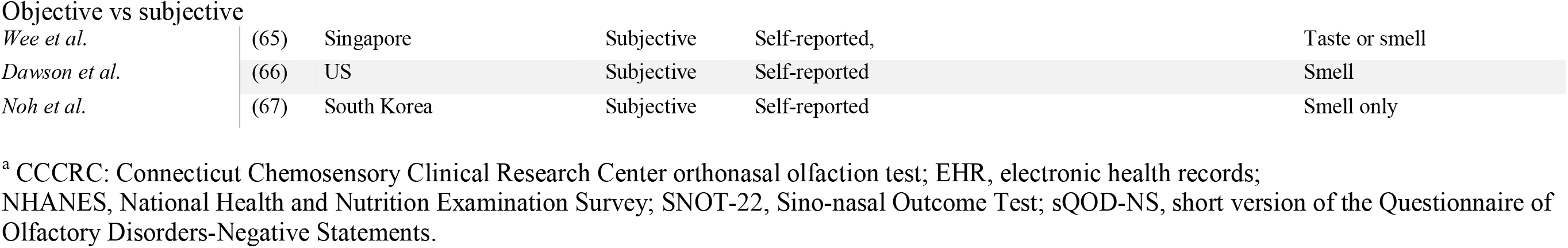
Summary of studies included in meta-analysis.

| *Article* | *Ref* | *Country* | *Subgroup* | *Specific sensory test<sup>a</sup>* | *Sense(s) measured* |
| --- | --- | --- | --- | --- | --- |
| *Vaira et al. 1* | (11) | Italy | Objective | CCCRC | Taste and smell, smell only |
| *Iravani et al.* | (5) | Sweden | Objective | Five-odor smell panel; used test-retest to measure reliability | Smell only |
| *Vaira et al. 2* | (12) | Italy | Objective | CCCRC, self-administered olfactory test | Smell only |
| *Vaira et al. 3* | (13) | Italy | Objective | CCCRC, home odor discrimination test | Smell |
| *Moein et al.* | (9) | Iran | Objective | University of Pennsylvania Smell Identification Test | Smell |
| *Hornuss et al.* | (16) | Germany | Objective | Sniffin' Sticks | Smell |
| *Parma et al.* | (50) | Global | Subjective | Self-reported | Smell |
| *Merza et al.* | (51) | Iraq | Subjective | Unknown, hospital reported | Smell only |
| *Levinson et al.* | (45) | Israel | Subjective | Self-reported | Smell only |
| *Haehner et al.* | (52) | Germany | Subjective | Self-reported | Smell only |
| *Speth et al.* | (53) | Switzerland | Subjective | Self-reported | Smell only |
| *De Maria et al.* | (54) | Italy | Subjective | Self-reported | Taste and smell |
| *Menni et al.* | (35) | UK and US | Subjective | Self-reported | Taste and smell |
| *Yan et al. 1* | (7) | US | Subjective | Self-reported | Smell only |
| *Luers et al.* | (55) | Germany | Subjective | Self-reported | Smell only |
| *Roland et al.* | (19) | US | Subjective | Self-reported | Taste or smell |
| *Boscolo-Rizzo et al.* | (56) | Italy | Subjective | Self-reported | Taste or smell |
| *Liu et al.* | (57) | Taiwan | Subjective | Unknown, hospital reported | Taste or smell |
| *Paderno et al.* | (21) | Italy | Subjective | Self-reported | Smell only |
| *Lee et al.* | (20) | Korea | Subjective | Self-reported | Taste or smell |
| *Lechien et al.* | (58) | Belgium, France, Spain, Italy | Subjective | Self-reported, survey based on NHANES and sQOD-NS | Smell only |
| *Gelardi et al.* | (59) | Italy | Subjective | Self-reported, | Taste and smell, smell only |
| *Giacomelli et al.* | (60) | Italy | Subjective | Self-reported | Taste and smell, smell only |
| *Shoer et al.* | (61) | Israel | Subjective | Self-reported | Taste or smell |
| *Mao et al.* | (8) | China | Subjective | Self-reported, EHR records | Smell only |
| *Spinato et al.* | (62) | Italy | Subjective | SNOT-22 | Taste or smell |
| *Beltran-Corbellini et al.* | (44) | Spain | Subjective | Self-reported | Smell only |
| *Trubiano et al.* | (17) | Australia | Subjective | Self-reported | Taste and smell, smell only |
| *Yan et al. 2* | (18) | US | Subjective | Self-reported | Smell |
| *Klopfenstein et al.* | (63) | France | Subjective | Self-reported | Smell |
| *Gudbjartsson et al.* | (64) | Iceland | Subjective | Self-reported | Taste or smell |
| *Wee et al.* | (65) | Singapore | Subjective | Self-reported, | Taste or smell |
| *Dawson et al.* | (66) | US | Subjective | Self-reported | Smell |
| *Noh et al.* | (67) | South Korea | Subjective | Self-reported | Smell only |

<sup>a</sup> CCCRC: Connecticut Chemosensory Clinical Research Center orthonasal olfaction test; EHR, electronic health records; NHANES, National Health and Nutrition Examination Survey; SNOT-22, Sino-nasal Outcome Test; sQOD-NS, short version of the Questionnaire of Olfactory Disorders-Negative Statements.

**Figure 2.**
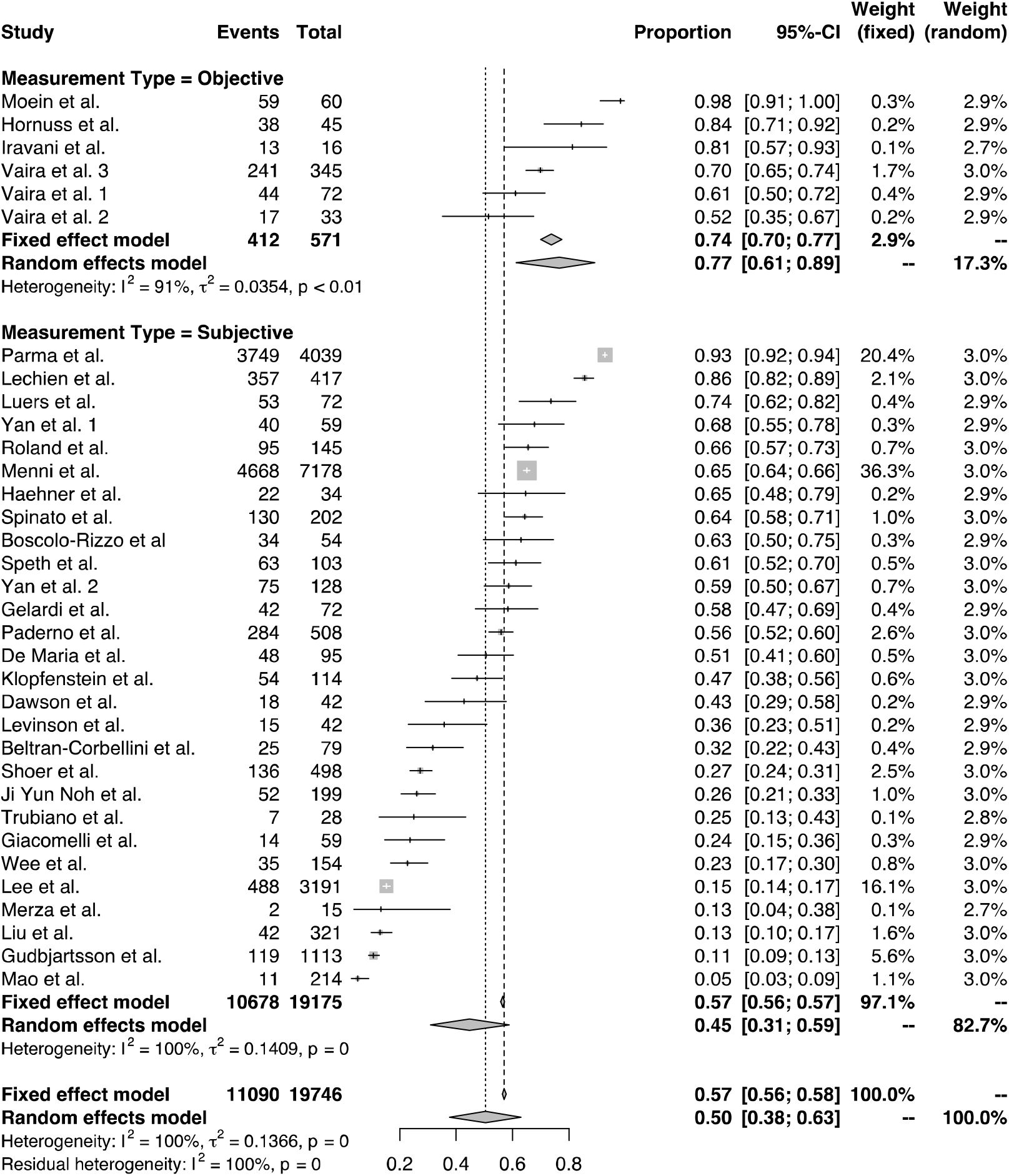
Forest plot meta-analysis of the prevalence of olfactory dysfunction in COVID-19 patients across studies classified as using objective (top) or subjective (bottom) methodologies. “Events” indicates cases of olfactory loss; “Total” indicates total number of COVID-19–positive patients. Both fixed-effects and random-effects models are presented. Individual study estimates are represented as “+” on the continuous horizontal line, which represents the 95% CI.

### Risk-of-Bias Assessment

Among the 34 studies included in this meta-analysis, none had a high risk of bias: 14 were categorized as low risk, and 20 as moderate risk. The risk-of-bias scores ranged from 2 to 6 across these studies, with an average risk of 3.79, indicating low to moderate risk of bias in our overall assessment.

### Prevalence of olfactory dysfunction in COVID-19 patients

Among the 34 studies, sample sizes ranged from 15 to 7,178 patients with positive verification of COVID-19. The number of cases of smell loss per study ranged from 2 to 4,668, with prevalence estimates ranging from 5% to 98.3%. Collectively, a total of 19,746 patients who tested positive for COVID-19 were included in this meta-analysis. Of these, 11,090 evidenced some form of olfactory dysfunction after infection with SARS-CoV-2. Meta-analysis for the pooled prevalence rate across all studies (*N*=34) yielded a significant Cochran’s *Q* (*Q*=8612.12, df=34, *p*<0.001) and *I*^2^ estimate of 99.6%. The pooled estimate for the prevalence rate for the overall cohort was 50.2% with a 95% CI of 37.7-62.6% (Figure 2).

### Effect of methodology on prevalence rate

Objective methods were used to assess olfactory loss in six studies, comprising 571 COVID-19 patients, with 412 reported cases of smell loss. Per study, the prevalence of olfactory loss ranged from 52% to 98.3% among COVID-19–positive patients. Pooled estimates of the prevalence rate were 73.9% and 76.7% under the fixed- and random-effect models, respectively. A significant Cochran’s *Q*, approximated from the chi-square distribution (*Q*=53.78, df=5, *p*<0.001), and *I*^2^ of 90.7% were obtained, confirming the heterogeneity of the data collected. When pooled across studies that utilized objective measurement tools, the average prevalence rate of olfactory loss is 76.7%, with a 95% CI of 61.4-89.2% (Figure 2).

A total of 28 studies were classified as using subjective methods (questionnaire, interview, etc.), comprising 19,175 subjects, with 10,678 cases of smell loss. The reported prevalence of olfactory loss ranged from 5% to 93% per study, a larger range than for studies classified as using objective methods. The pooled estimates of the prevalence rate were 56.49% and 44.58% under the fixed- and random-effect models, respectively. Similar to the objective subgroup, Cochran’s *Q* was significant (*Q*=8487.92, df=28, *p*<0.001), and the *I*^2^ value was 99.7%, confirming the heterogeneity of the meta-analysis across the subjective studies. To account for the observed heterogeneity, we report the prevalence rate of 44.6% estimated by the random-effects model, with a 95% CI of 31.1-58.5% (Figure 2).

## Discussion

This systematic review and meta-analysis revealed that olfactory dysfunction is a prominent symptom of COVID-19. Meta-analysis using the random-effects model computed an overall prevalence rate of 50.2% (95% CI: 37.7-62.6%), which is very similar to, although slightly lower than, the previously reported value of 53% in a meta-analysis of olfactory dysfunction in 10 studies (23). Both meta-analyses confirm that olfactory dysfunction, regardless of the measurement methodology, is identified in about half of the patients infected with SARS-CoV-2.

### Methodological differences in smell loss measurement tools impact reported prevalence rate of olfactory loss in COVID-19 patients

We further examined whether there were differences in prevalence of olfactory loss based on the type of method used to gather such information. Most studies (28 of 34) included in this meta-analysis were classified as using subjective methods (self-report) to quantify the prevalence of smell loss; 6 studies used objective methods (e.g., odor threshold tests). The reliance on self-report was expedient due to the pandemic conditions and global stay-at-home orders. However, our analysis revealed a stark difference in prevalence between the two subgroups; studies using objective methods reported around 77% prevalence overall, whereas those using subjective methods reported around 45%.

There are inherent pros and cons regarding each type of methodology. Objective methods quantify smell loss and can limit any confounds because they are often conducted in a controlled environment with standardized procedures. Objective methods rely on true perception of a stimuli when presented, diminishing response and measurement bias. In contrast, subjective methods naturally encompass more variability due to a lack of standardization in how and what questions were asked. Additionally, subjective methods are often prone to recall bias. However, they are an easy and cost-efficient way to collect information quickly from the intended population, as demonstrated by the numerous studies in our meta-analysis that used this type of method. Objective methods have higher time and cost requirements than do subjective methods.

The higher overall reported prevalence of olfactory loss in studies using objective methods (77%) compared to those using subjective methods (45%) suggests that subjective methodologies miss crucial information and might consistently underestimate true smell loss in COVID-19 patients. One major difference between the two types of methods is the number of variables assessed in each study. Often in the studies using subjective methods the researchers were interested in numerous aspects of COVID-19 symptoms, not just smell loss alone, whereas the studies using objective methods focused solely on sensory loss, using numerous stimuli to get a sensitive measurement of the patient’s smell loss. Our findings align with a prior meta-analysis by Tong et al. (23) that found that non-standardized methods (which were all subjective methods by our criteria) severely underestimated olfactory prevalence (estimated at ∼37%) compared to standardized methods (which included both subjective and objective methods), which indicated around 87% prevalence. Standardized methods outlined in the Tong et al. study consisted of both objective and subjective measures—because a method is subjective does not mean that the method is not standardized or validated. Validated subjective methods for collecting information on olfactory dysfunction in our pool of studies include a version of the Questionnaire of Olfactory Dysfunction, and the Sino-Nasal Outcome Test (42, 43).

Overall, however, objective methods have been found to be more sensitive in detecting anosmia and hyposmia than subjective self-reports in a COVID-19 patient population (16). Additionally, among patients who initially self-reported no smell loss, objective analysis showed mild hyposmia in 30%, again pointing to underreporting by subjective methods (13). On the other hand, in a COVID-19 patient population recruited due to suspected olfactory loss, 38% of patients with self-reported olfactory dysfunction had normal olfactory performance using the Sniffin’ Sticks test, an objective method (15). This overreporting could be due to the biased and specific recruitment (patients with suspected olfactory loss) compared to the general COVID-19 patient population recruited for studies in our meta-analysis.

The higher reported prevalence of olfactory loss when using objective as compared to subjective methods to measure olfactory loss calls for further examination of the consequences of the methodologies employed. Researchers might be missing a critical symptom of COVID-19 through the use of unstandardized, subjective methods to measure smell loss, as demonstrated by the lower prevalence rate we found in studies classified as using subjective methods. However, objective methods are costly and time-consuming to conduct in standardized laboratory settings.

Many researchers have adapted objective methods to evaluate smell loss to enable use in a home setting. Varia et al found no significant difference in patient smell loss ratings when conducted with an objective method during hospitalization (standardized setting) versus during home quarantine (13). Irvani et al. used an app to track smell loss in COVID-19 patients over time, which revealed moderate test-retest reliability across sessions among users showing no symptoms, and significant reduction in olfactory function for those who tested positive for COVID-19 compared to those who tested negative (5). The accessibility and adaptability of these objective approaches make them a resource-efficient strategy to obtain an accurate measure of olfactory loss in COVID-19 patients.

Anosmia is frequently reported to be one of the first presenting symptoms of COVID-19 (23, 44, 45). In a cohort specifically of patients complaining of smell loss, researchers found that 83% of people reported anosmia as their first symptom of COVID-19 (32). The actual mechanism by which SARS-CoV-2 may inhibit and disrupt smell perception is currently unknown; however, many potential theories about the cause of smell loss have been proposed. Reports suggest that, different from other coronaviruses, such as those that cause the common cold, SARS-CoV-2 can cause smell loss even in the absence of symptoms such as blockage of the nose, postnasal drip, or a runny nose, which are typical co-occurring manifestations of smell loss from other respiratory viruses (46). The lack of nasal blockage suggests COVID-19 might be a neurotropic and neuroinvasive disease (47). Furthermore, it is now commonly known that SARS-CoV-2 coronavirus binds to ACE2 receptors, allowing the virus to enter and infect cells. ACE2 receptors are expressed in nasal epithelium cells (48), specifically the structures that support olfactory neurons, leading to a theory that infection of these supporting cells might cause additional damage to the olfactory epithelium, resulting anosmia or hyposmia (49).

Though olfactory dysfunction occurs in high prevalence rates in patients positive for COVID-19, time to recovery varies across the studies. Several studies mention significant improvements quickly after symptom onset, e.g., (7). Other studies reported that many patients still had not returned to normal sense of smell more than 2 weeks after initial onset of smell loss (16). Altogether, our meta-analysis demonstrates a prevalence of identified smell loss in about half of the COVID-19 patients, supporting the need to understand the mechanism of infection, onset of symptoms, and recovery from olfactory loss due to a SARS-CoV-2 infection.

### Limitations and Future Research

Due to the nature of data collection amidst an evolving global pandemic, there are inherent limitations to the present meta-analysis, many of which were driving factors of the observed high heterogeneity across studies. Disease severity of the recruited study population (COVID-19–positive patients) was not controlled for, which could add to selection bias. A wide range of measurement methods were employed within both objective and subjective categories, which naturally creates measurement bias. Often recall bias occurs in subjective methodologies, as self-recognition may occur only in severe cases and is often forgotten in prolonged, more subtle cases (47). Furthermore, there is lack of awareness regarding chemosensory function in subjects—many researchers combined the “loss of taste or smell” in their symptomatic findings, even though they are two completely different perceptions that would be impacted differently by SARS-CoV-2. In addition, there remains a lack of comprehensive testing of chemesthetic sensations (e.g., burn from capsaicin or cooling from menthol compounds) (50).

Assessment of olfactory function in patients with suspected or confirmed COVID-19 diagnosis may become standard practice by clinicians. Despite the limitations inherit in subjective measures, at a minimum, patients need to be interviewed about their sense of smell as a first-line assessment. Given the interrelationship between smell and taste, during clinical assessments patients may report changes in taste rather than changes in smell. For patients who report changes in smell and taste function during screening questionnaires, full testing should be performed using objective standardized chemosensory assessment tools. Considering that psychophysical testing may not be possible for all patients and the current social distancing regulations, regular olfactory and gustatory self-assessment at home may be an initial recommendation. Although regular self-assessment may give information about chemosensory function during the trajectory of the disease, the results should be interpreted with caution. In addition, longitudinal assessments of chemosensory function may help identify those patients with continued impairment who may need further treatment and non-pharmacological interventions (e.g., olfactory training).

More research is needed to better establish procedures to estimate prevalence rates of sensory loss. Our meta-analysis results reveal underestimates when using subjective techniques, supporting the value of adapting objective methods to estimate smell loss. As information regarding COVID-19 is constantly evolving and is being crowd-sourced, more than ever researchers need to come together on methods to best assess smell loss.

## Data Availability

All code and data are available on Github

https://github.com/vramirez4/OlfactoryLoss

## Acknowledgements

We would like to acknowledge Michael G. Tordoff for his assistance with the review of the literature. Input from the members of the Global Consortium for Chemosensory Research including Valentina Parma, Kathrin Ohla, Thomas Hummel, Steven Munger, John Hayes, Chrissi Kelly, Marga Veldhuizen, and Masha Niv is acknowledged. We thank William Kyle Hamilton and Beverly Cowart for their comments on this manuscript. A companion website updated frequently with new study results is available at https://vicente-ramirez.shinyapps.io/COVID19_Olfactory_Dashboard/.

---

**Supplementary Table S1.**
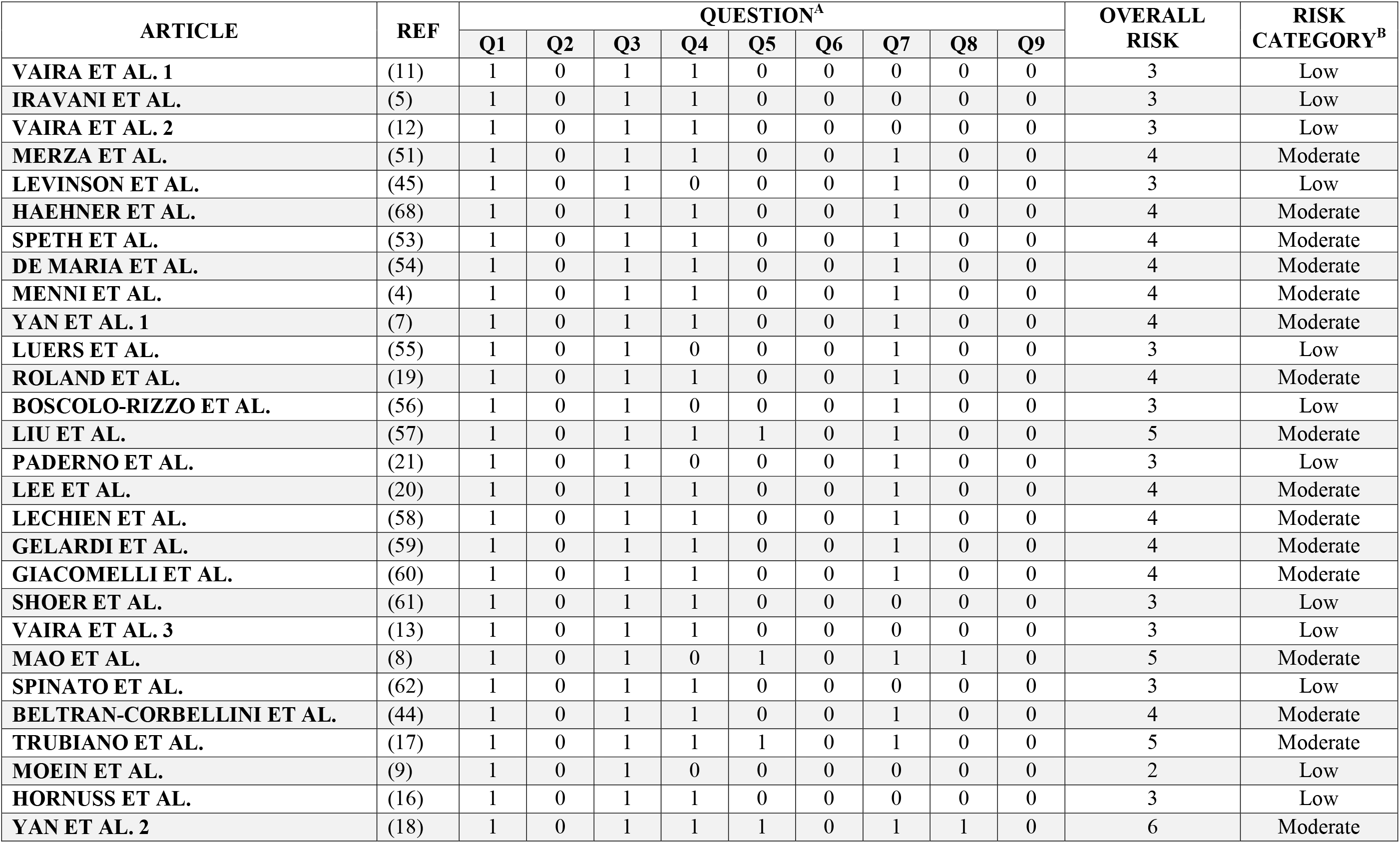
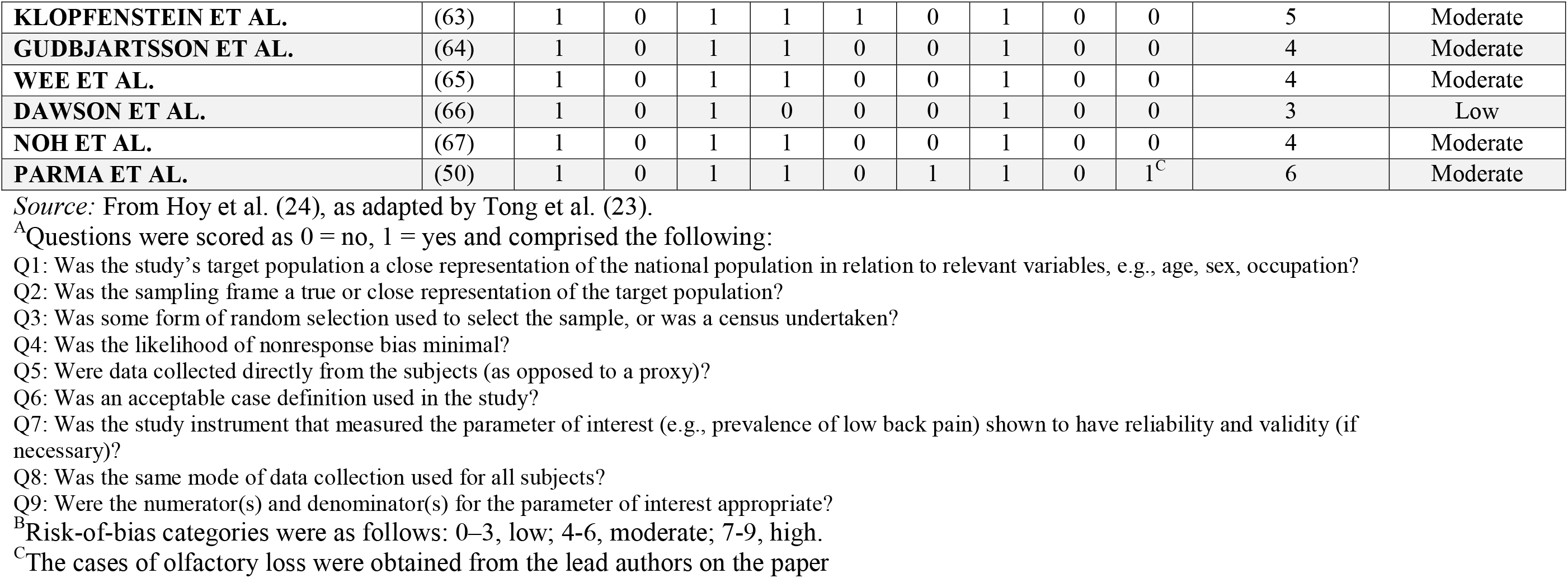
Risk-of-bias assessment of selected articles.

| ARTICLE | REF | QUESTION<sup>A</sup> |  |  |  |  |  |  |  |  | OVERALL RISK | RISK CATEGORY<sup>B</sup> |
| --- | --- | --- | --- | --- | --- | --- | --- | --- | --- | --- | --- | --- |
|  |  | Q1 | Q2 | Q3 | Q4 | Q5 | Q6 | Q7 | Q8 | Q9 |  |  |
| VAIRA ET AL. 1 | (11) | 1 | 0 | 1 | 1 | 0 | 0 | 0 | 0 | 0 | 3 | Low |
| IRAVANI ET AL. | (5) | 1 | 0 | 1 | 1 | 0 | 0 | 0 | 0 | 0 | 3 | Low |
| VAIRA ET AL. 2 | (12) | 1 | 0 | 1 | 1 | 0 | 0 | 0 | 0 | 0 | 3 | Low |
| MERZA ET AL. | (51) | 1 | 0 | 1 | 1 | 0 | 0 | 1 | 0 | 0 | 4 | Moderate |
| LEVINSON ET AL. | (45) | 1 | 0 | 1 | 0 | 0 | 0 | 1 | 0 | 0 | 3 | Low |
| HAEHNER ET AL. | (68) | 1 | 0 | 1 | 1 | 0 | 0 | 1 | 0 | 0 | 4 | Moderate |
| SPETH ET AL. | (53) | 1 | 0 | 1 | 1 | 0 | 0 | 1 | 0 | 0 | 4 | Moderate |
| DE MARIA ET AL. | (54) | 1 | 0 | 1 | 1 | 0 | 0 | 1 | 0 | 0 | 4 | Moderate |
| MENNI ET AL. | (4) | 1 | 0 | 1 | 1 | 0 | 0 | 1 | 0 | 0 | 4 | Moderate |
| YAN ET AL. 1 | (7) | 1 | 0 | 1 | 1 | 0 | 0 | 1 | 0 | 0 | 4 | Moderate |
| LUERS ET AL. | (55) | 1 | 0 | 1 | 0 | 0 | 0 | 1 | 0 | 0 | 3 | Low |
| ROLAND ET AL. | (19) | 1 | 0 | 1 | 1 | 0 | 0 | 1 | 0 | 0 | 4 | Moderate |
| BOSCOLO-RIZZO ET AL. | (56) | 1 | 0 | 1 | 0 | 0 | 0 | 1 | 0 | 0 | 3 | Low |
| LIU ET AL. | (57) | 1 | 0 | 1 | 1 | 1 | 0 | 1 | 0 | 0 | 5 | Moderate |
| PADERNO ET AL. | (21) | 1 | 0 | 1 | 0 | 0 | 0 | 1 | 0 | 0 | 3 | Low |
| LEE ET AL. | (20) | 1 | 0 | 1 | 1 | 0 | 0 | 1 | 0 | 0 | 4 | Moderate |
| LECHIEN ET AL. | (58) | 1 | 0 | 1 | 1 | 0 | 0 | 1 | 0 | 0 | 4 | Moderate |
| GELARDI ET AL. | (59) | 1 | 0 | 1 | 1 | 0 | 0 | 1 | 0 | 0 | 4 | Moderate |
| GIACOMELLI ET AL. | (60) | 1 | 0 | 1 | 1 | 0 | 0 | 1 | 0 | 0 | 4 | Moderate |
| SHOER ET AL. | (61) | 1 | 0 | 1 | 1 | 0 | 0 | 0 | 0 | 0 | 3 | Low |
| VAIRA ET AL. 3 | (13) | 1 | 0 | 1 | 1 | 0 | 0 | 0 | 0 | 0 | 3 | Low |
| MAO ET AL. | (8) | 1 | 0 | 1 | 0 | 1 | 0 | 1 | 1 | 0 | 5 | Moderate |
| SPINATO ET AL. | (62) | 1 | 0 | 1 | 1 | 0 | 0 | 0 | 0 | 0 | 3 | Low |
| BELTRAN-CORBELLINI ET AL. | (44) | 1 | 0 | 1 | 1 | 0 | 0 | 1 | 0 | 0 | 4 | Moderate |
| TRUBIANO ET AL. | (17) | 1 | 0 | 1 | 1 | 1 | 0 | 1 | 0 | 0 | 5 | Moderate |
| MOEIN ET AL. | (9) | 1 | 0 | 1 | 0 | 0 | 0 | 0 | 0 | 0 | 2 | Low |
| HORNUSS ET AL. | (16) | 1 | 0 | 1 | 1 | 0 | 0 | 0 | 0 | 0 | 3 | Low |
| YAN ET AL. 2 | (18) | 1 | 0 | 1 | 1 | 1 | 0 | 1 | 1 | 0 | 6 | Moderate |
| **KLOPFENSTEIN ET AL.** | (63) | 1 | 0 | 1 | 1 | 1 | 0 | 1 | 0 | 0 | 5 | Moderate |
| **GUDBJARTSSON ET AL.** | (64) | 1 | 0 | 1 | 1 | 0 | 0 | 1 | 0 | 0 | 4 | Moderate |
| **WEE ET AL.** | (65) | 1 | 0 | 1 | 1 | 0 | 0 | 1 | 0 | 0 | 4 | Moderate |
| **DAWSON ET AL.** | (66) | 1 | 0 | 1 | 0 | 0 | 0 | 1 | 0 | 0 | 3 | Low |
| **NOH ET AL.** | (67) | 1 | 0 | 1 | 1 | 0 | 0 | 1 | 0 | 0 | 4 | Moderate |
| **PARMA ET AL.** | (50) | 1 | 0 | 1 | 1 | 0 | 1 | 1 | 0 | 1<sup>C</sup> | 6 | Moderate |

*Source:* From Hoy et al. (24), as adapted by Tong et al. (23).
<sup>A</sup>Questions were scored as 0 = no, 1 = yes and comprised the following:
Q1: Was the study's target population a close representation of the national population in relation to relevant variables, e.g., age, sex, occupation?
Q2: Was the sampling frame a true or close representation of the target population?
Q3: Was some form of random selection used to select the sample, or was a census undertaken?
Q4: Was the likelihood of nonresponse bias minimal?
Q5: Were data collected directly from the subjects (as opposed to a proxy)?
Q6: Was an acceptable case definition used in the study?
Q7: Was the study instrument that measured the parameter of interest (e.g., prevalence of low back pain) shown to have reliability and validity (if necessary)?
Q8: Was the same mode of data collection used for all subjects?
Q9: Were the numerator(s) and denominator(s) for the parameter of interest appropriate?
<sup>B</sup>Risk-of-bias categories were as follows: 0–3, low; 4–6, moderate; 7–9, high.
<sup>C</sup>The cases of olfactory loss were obtained from the lead authors on the paper

